# Epidemiological characteristics of SARS-COV-2 in Myanmar

**DOI:** 10.1101/2020.08.02.20166504

**Authors:** Aung Minn Thway, Htun Teza, Tun Tun Win, Ye Min Htun, Moe Myint Aung, Yan Naung Win, Kyaw Myo Tun

## Abstract

Coronavirus disease (COVID-19) is an infectious disease caused by a newly discovered severe acute respiratory syndrome coronavirus 2 (SARS-CoV-2). In Myanmar, first COVID-19 reported cases were identified on 23^rd^ March 2020. There were 336 reported confirmed cases, 261 recovered and 6 deaths through 13^th^ July 2020. The study was a retrospective case series and all COVID-19 confirmed cases from 23^rd^ March to 13^th^ July 2020 were included. The data series of COVID-19 cases were extracted from the daily official reports of the Ministry of Health and Sports (MOHS), Myanmar and Centers for Disease Control and Prevention (CDC), Myanmar. Among 336 confirmed cases, there were 169 cases with reported transmission events. The median serial interval was 4 days (IQR 3, 2-5) with the range of 0 - 26 days. The mean of the reproduction number was 1.44 with (95% CI = 1.30-1.60) by exponential growth method and 1.32 with (95% CI = 0.98-1.73) confident interval by maximum likelihood method. This study outlined the epidemiological characteristics and epidemic parameters of COVID-19 in Myanmar. The estimation parameters in this study can be comparable with other studies and variability of these parameters can be considered when implementing disease control strategy in Myanmar.

## Introduction

Coronavirus disease (COVID-19) is an infectious disease caused by a newly discovered severe acute respiratory syndrome coronavirus 2 (SARS-CoV-2). It was first identified as the cause of a cluster of pneumonia cases in Wuhan, a city in the Hubei Province of China at the end of 2019, and then it was rapidly spreading in China and other countries [1, 2]. Consequently, COVID-19 has become a major global health concern, and it was recognized as a pandemic by the World Health Organization (WHO) on 11^th^ March 2020 [3]. The daily total number of cases were steady rising with affecting 213 countries and territories around the world. Globally, there were more than 12 million (12,768,307) confirmed cases including 566,654 deaths through 13^th^ July 2020 [4]. United States of America, Brazil, India, Russia Federation and Peru were the most effected countries. Among WHO regions, America, Europe and Eastern Mediterranean Regions were highest affected regions. In South East Asia region, there were more than one million (1,163,556) confirmed cases and 31,297 case are deaths [4, 5].

SARS-CoV-2 can be spread by human-to-human transmission via droplets or direct contact [6]. The infection has been estimated to have mean incubation period of 6.4 days and a mean basic reproduction number (*R*_0_) was 3.38 with a range of 1.90 to 6.49 [7-9]. The incubation period for COVID-19 is on average 5-6 days, but it can be up to 14 days [10]. Moreover, as outliers, the patients with incubation period of 19 days [11] and 24 days [12] have been reported with potential asymptomatic transmission. Even though there is no clear evidence as yet of asymptomatic transmission, the relatively high proportion of asymptomatic infections could have public health implications [13].

There were estimated mean serial interval for COVID-19 of 3.96 days [14], 4.4 days [15], 6.5 days [16], 4.7 days [17] and 4.8 days [18]. It was close to or shorter than its median incubation period, and a substantial proportion of secondary transmission might occur prior to illness onset [17]. Most people with COVID-19 would experience mild to moderate respiratory illness and recover without requiring special treatment. However, older people, and those with underlying medical problems such as cardiovascular disease, diabetes, chronic respiratory disease, and cancer were more likely to develop serious illness [1].

In Myanmar, first COVID-19 reported cases were identified on 23^rd^ March 2020. There were 336 reported confirmed cases, 261 recovered and 6 deaths through 13^th^ July 2020. Obviously, reported confirmed cases were gradually increased in Yangon Region, an epicenter of COVID-19 outbreak, and majority of cases (242 cases) were reported from this region [19]. The government of Myanmar has extended several restrictions including the suspension of international commercial flights, bans on public gatherings, and closures of public events, entertainment venues and religious institutions. Since 16^th^ May 2020, almost new cases were imported cases who returned back from other countries and they have already been in quarantine centres. Most cases were identified in people return back from foreign countries [20]. The COVID-19 pandemic has disrupted economic activities and is expected to have a long-term impact on various sectors.

The critical importance are epidemiologic investigations to characterize mode of transmission, reproduction number, serial interval, and clinical spectrum of infection in order to reform and refine strategies that can stop the spread of COVID-19 [21]. The characterization of the epidemiological features of COVID-19 is also vital for the development and implementation of effective prevention and control measures. This study aimed to describe the epidemiology characteristics and main epidemiological parameters of COVID-19 in Myanmar.

## Methods

### Study design and population

The study was a retrospective case series and all COVID-19 confirmed cases from 23^rd^ March to 13^th^ July 2020 were included.

### Data retrieval

The data series of COVID-19 cases were extracted from the daily official reports of the Ministry of Health and Sports (MOHS), Myanmar [20, 22] and Centers for Disease Control and Prevention (CDC), Myanmar [23]. Publicly accessible data including total tested, new and cumulative number of confirmed cases, negative cases, recovered cases, active cases and deaths, and the closed contacts are updated daily by the MOHS [20, 22] and CDC, Myanmar [23]. The epidemiological characteristics of COVID-19 cases were age, sex, geographical distribution, date of symptom onset, date of admission, date of report, travel history, contact history, number of contacts and type of transmission were retrieved. However, date of symptom onset cannot not be accessible form the data sources, therefore the date of test positive of confirmed cases is assumed as such. A confirmed case of COVID-19 infection was defined as a case with a positive result for viral nucleic acid testing, real-time RT-PCR, in respiratory specimens [24].

### Statistical analysis

Descriptive analyses of the variables were expressed as mean (± Standard Deviation, SD), median with (Interquartile Range, IQR) and number (%). The distributions of age and sex, epidemic curve and distribution of estimated serial interval were shown by GraphPad Prism version 7(GraphPad Software). The mean of basic reproductive number with 95 % Confidence Interval (CI) was calculated. The data were analyzed using Excel version 2016, *R*_0_ package (version 1.2-6 released in May 2015) and Epicontacts package (version 1.1.1 released in May 2017) of *R* environment (version 3.6.2 released in December 2019). The data were analyzed using Excel version 2016, *R*_0_ package (version 1.2-6 released in May 2015) and Epicontacts package (version 1.1.1 released in May 2017) of *R* environment (version 3.6.2 released in December 2019).

### Outcomes

The Epidemiological parameter included epidemic curve, basic reproduction number (*R*_0_), serial interval and close contact of a confirmed case. An epidemic curve is a visual display of the onset of illness among cases associated with an outbreak. The general sense of the outbreak’s magnitude and inferences about the outbreak’s pattern of spread could be estimated [25]. *R*_0_ was defined as the expected number of secondary cases that one primary case will generate in a susceptible population. If *R*_0_ is above the critical threshold of 1, continuous human-to-human transmission with sustained transmission will occur [24]. *R*_0_ package implements a simple, likelihood-based estimation of *R*_0_ using a branching process with a Poisson likelihood. Infectiousness is determined by weighting *R*_0_ by the probability mass function of the serial interval on the corresponding day. It is a simplified version of the model introduced by Jombart T et al. in 2013 [26]. Reproduction numbers may be calculated by different method. The exponential method and maximum likelihood method were used to estimate the reproduction numbers in this study.

The serial interval of COVID 19 is defined as the time elapsed between a primary case-patient (infector) having symptom onset and a secondary case-patient (infectee) having symptom onset [24]. Regarding epidemiological contact with confirmed case, contact plot was created by using Epicontacts. It is an *R*^15^ package providing a suite of tools aimed at merging line lists and contact data, and providing basic functionality for handling, visualizing and analyzing epidemiological contact data. Maintained as part of the *R* Epidemics Consortium (RECON), this package is integrated into an ecosystem of tools for outbreak response using the R language.

## Results

A total of 336 confirmed cases, 261 recovered cases, 69 active cases and 6 deaths were reported from 23^rd^ March to 13^th^ July 2020. All confirmed cases were referred to and treated in designated hospital of respective States and Regions.

### Epidemiological characteristics

Based on reported data, mean (± SD) age of COVID-19 confirmed cases was 36 (± 15.89) years and so, working age groups were more contracted to infection in both sexes although all age groups could be infected. As shown in Fig. 1, males were slightly more risk to get infection compared to females.

**Figure 1.**
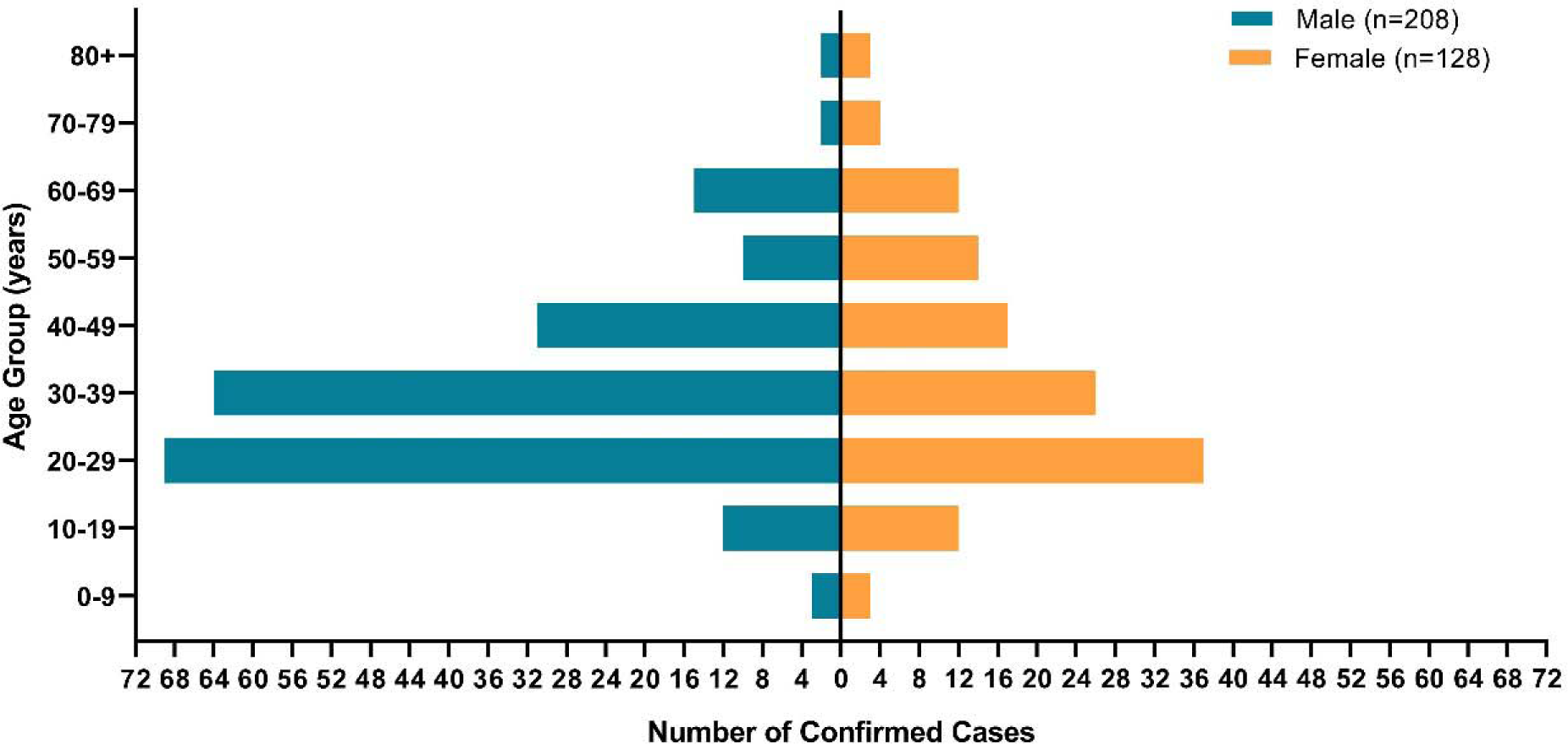
Age and sex distribution of confirmed COVID-19 cases in Myanmar through 13^th^ July 2020 (n=336)

On place of acquisition over time (Fig. 2), imported cases are 176 (52.38%), local transmission cases with close contact are 142 (42.26%) and local transmission cases with no epidemiological linked are 18 (5.36%) respectively.

**Figure 2.**
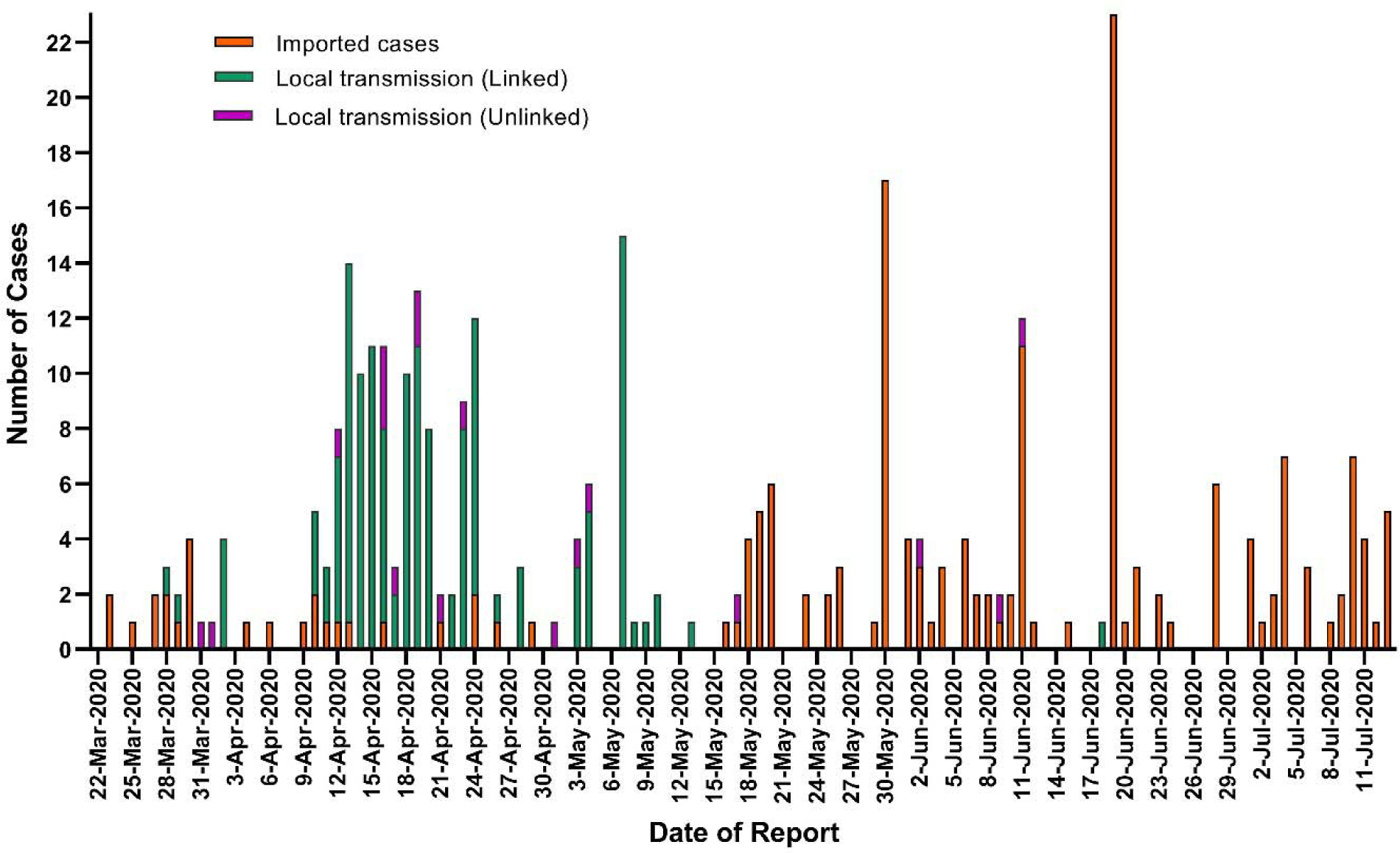
Epidemiological curve of COVID-19 in Myanmar through 13^th^ July 2020, showing with imported cases (orange), local transmission with close contact (green), and local transmission with no epi-linked (purple).

As a geographical distribution, among fifteen States and Regions, the majority of cases have been identified in Yangon Region (242, 72.02%), Kayin State (26, 7.74%) and Rakhine State (14, 4.17%). Only one State (Kayah) has no report of COVID-19 confirmed cases through 13th July 2020.

### Epidemiological parameters

Among 336 confirmed cases, there were 169 cases with reported transmission events. The median serial interval was 4 days (IQR 3, 2-5) with the range of 0 - 26 days as shown in Fig. 3.

**Figure 3.**
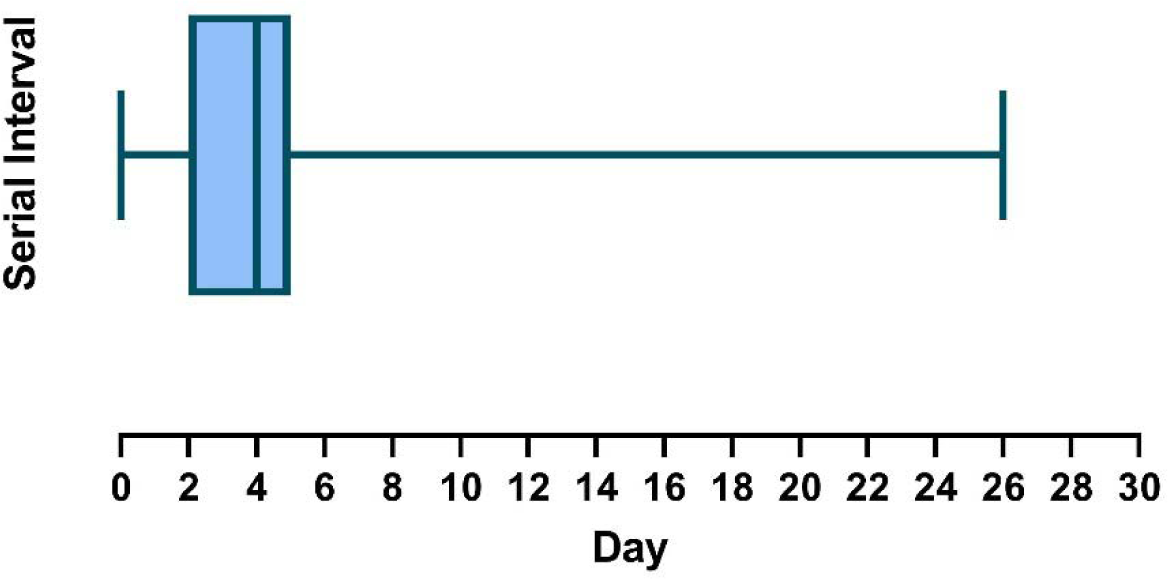
Estimated serial interval distribution for COVID-19 confirmed cases with reported transmission events in Myanmar through 13^th^ July 2020

**Figure 4.**
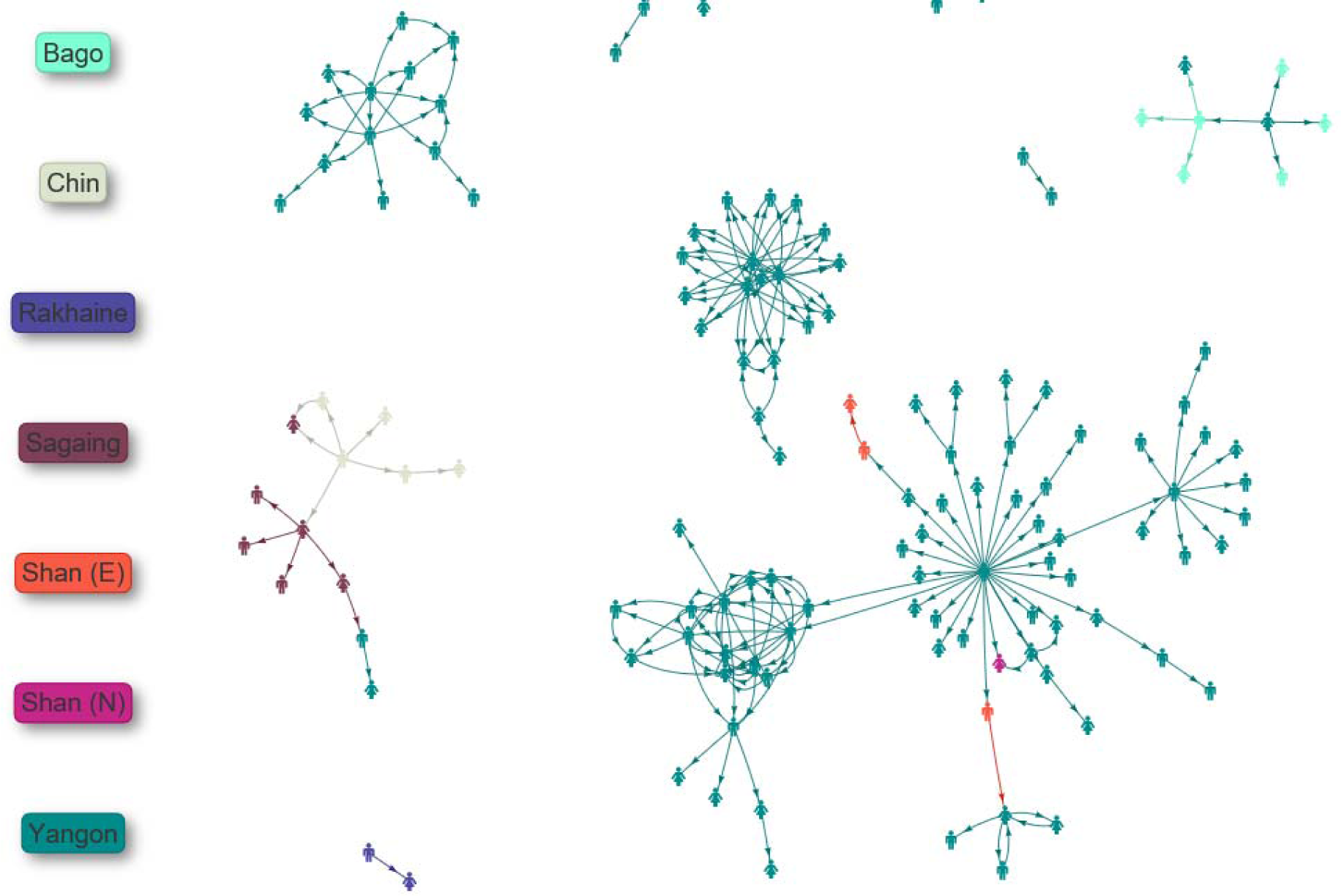
Contact plot for COVID-19 confirmed cases with reported transmission events in Myanmar through 13^th^ July 2020.

For 336 confirmed cases through 13^th^ July 2020, the mean of the reproduction number was 1.44 with (95% CI = 1.30-1.60) by exponential growth method and 1.32 with (95% CI = 0.98-1.73) confident interval by maximum likelihood method (Table 1).

**Table (1).**
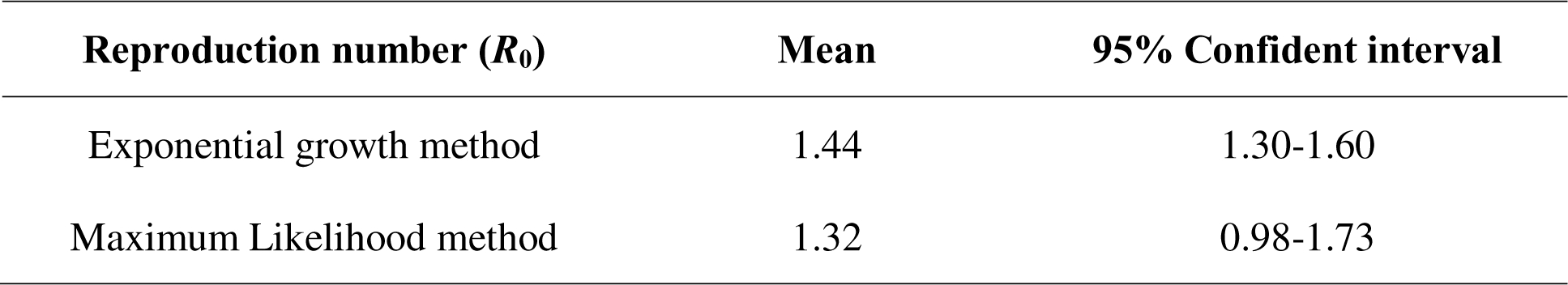
Estimated basic reproduction number at 13^th^ July 2020.

| Reproduction number ($R_0$) | Mean | 95% Confident interval |
| --- | --- | --- |
| Exponential growth method | 1.44 | 1.30-1.60 |
| Maximum Likelihood method | 1.32 | 0.98-1.73 |

With regard epidemiological contact, 4 clusters of cases were obviously detected in Myanmar. The first cluster of cases was originated from case-1 to additional (11) confirmed cases out of (137) contact persons. The second cluster of cases was emerged from case-8,11,12,13 to additional (15) confirmed cases out of (714) contact persons. The third cluster of cases was contributed to highest proportion of confirmed cases. It was originated from case-24 to additional (78) confirmed cases among (396) contact persons. The last cluster of cases was originated from case-162 to additional (20) confirmed cases.

## Discussion

This study set out to provide epidemiological characteristics and main epidemiological parameters of COVID-19 in Myanmar. Although most cases (242 out of 336 cases) were detected in Yangon Region, the disease has spread to the whole country except one out of fifteen states, regions and union territory within 2 months. Myanmar is one of the least affected countries of COVID-19 in South East Asian Region. Current basic reproduction number (*R*_0_) and serial interval was estimated based on available reported data. Unfortunately, the whole-genome sequences of SARS-CoV-2 isolated from confirmed cases in Myanmar is currently unavailable and the strain of coronavirus could not also identify. Since detection of first case of COVID-19 in Myanmar, case fatality rate was 1.79% (6 out of 366). In contrast to mortality rate in South East Asia Region (2.69%) and world population (4.34%), disease mortality was substantially lower [4, 5]. The observed decrease in case fatality rate in Myanmar could be attributed to all laboratory-confirmed cases were admitted to designated specialist hospitals and continuous monitoring of cases for two consecutive negative results during hospitalization.

By 13^th^ July 2020, Myanmar had reported 336 cases, but the outbreak trajectory has been slow compared with neighboring countries such as India, China, Bangladesh and Thailand. This could be attributed to early implementation of non-pharmaceutical interventions. On 13^th^ March 2020, the President Office of Myanmar announced on avoidance of mass gathering followed by school closure on 20^th^ March 2020. Hand washing campaign was encouraged by State Counselor on 21^st^ March 2020 and social distancing campaign on 25^th^ March 2020. On 29^th^ March 2020, Myanmar has decided to adopt the restrictions for visitors from all countries by the temporary suspension of the issuance of all types of visa except diplomats accredited to Myanmar, United Nations official residents in Myanmar and crew of ships and aircraft operating to and from Myanmar. MOHS announced stay at home order and lock down by enforcing law on 10^th^ April 2020 and 18^th^ April 2020.

Myanmar is one of the low income countries and resources for diagnostic capacities were limited to compare with developed countries. However, daily test performance was less than one hundred initially, it is significantly growth in testing capacity to more than one thousand per day in recent time. The diagnosis is now focused on people returned back from other countries since mid-May 2020 because the importation of COVID-19 to Myanmar was greatly contributed to daily detection of confirmed cases. So the imported cases were 52.38% of total confirmed cases in Myanmar. Government planned many quarantine centers throughout the whole countries for those people and there is no exception for quarantine. This activity is an important strength in containment of COVID-19 infection in Myanmar.

In this study, two key epidemic parameters were estimated for further spread of COVID-19 in Myanmar. Firstly, the basic reproduction number *R*_0_ was 1.44 with (95% CI = 1.30-1.60) by exponential growth method and 1.32 with (95% CI = 0.98-1.73) confident interval by maximum likelihood method by using the real-time reports on the number of COVID-19 cases from the daily official reports of MOHS. In contrast to preliminary prediction in China, the basic reproduction number in Myanmar was substantially lower than China whereas *R*_0_ was fallen between 2.8 and 3.3 [27], and Diamond Princess Cruise ship whereas *R*_0_ was about 2.28 (2.06 – 2.52) [28]. Recently, *R*_0_ in Myanmar is approximately at 1.3 - 1.4 and, so it can be assumed that the outbreak will be continued slowly in nature. But, there are no local transmission cases since mid-May 2020 and the prediction of basic reproduction number could not be effective in current situation.

Secondly, serial interval, the time elapsed between symptom onsets of two successive generations of cases. The median serial interval was 4 days (IQR 3, 2-5) with the range of 0 - 26 days and this estimation was based on 169 cases with reported transmission events. The range of 0-26 days may be explained by the fact that reported contact history by patients and some contact tracing may be lack of detailed information. The findings of current study are consistent with serial interval in Hong Kong, mean serial interval was 4.58 days (95% CI: 3.35 to 5.85) but it is slightly differ from Li et al., it was a mean serial interval of 7.5 days (95% CI, 5.3 to 19) [29, 30]. The longest serial interval was observed about 26 days in Myanmar and hence, 28-days quarantine strategy of MOHS is well covered for all contacts with suspected cases.

This study is an epidemiological characteristic of COVID-19 cases in Myanmar, and we recognize the following limitations. First, the actual cases in the country within the study period could be underestimated due to low testing capabilities. Second, the recall bias of cases might have affected data accuracy because of self-reported contact history and symptom onset. Third, with slight inaccuracies of epidemiological information on confirmed cases reported by MOHS, more detailed information regarding date of symptom onset and clinical features were unavailable at the time of analysis. Therefore, dates of test positive of confirmed cases were inferred as the dates of symptom onset to explore the serial interval of transmission event. The closed contacts were identified within a few days after confirmation of their contact history, and were kept at quarantined centers till the result came out. It could be expected that the variation of this duration would be minimal. Fourth, for some patients, the official reports link the person with multiple confirmed cases as contacts. This is not a problem unique to our study but in the other similar studies as well, especially in clusters. This is addressed by considering the earliest confirmed case of the contacts as the source.

## Conclusion

This study outlined the epidemiological characteristics and epidemic parameters of COVID-19 in Myanmar. The estimation parameters in this study can be comparable with previous documents by others and variability of these parameters can be considered when implementing disease control strategy. Most probably Myanmar is now at mitigation phase because we rarely found on local transmission case in our country. In order to suppress this outbreak, government must be strengthened current successful non-pharmaceutical interventions, maintaining of non-COVID-19 essential health services and maintaining of essential societal services.

## Data Availability

All data are retrieved from Myanmar Ministry of Health and Sports.

